# Elevated expression of *RGS2* may underlie reduced olfaction in COVID-19 patients

**DOI:** 10.1101/2022.05.12.22274991

**Authors:** Eden Avnat, Guy Shapira, David Gurwitz, Noam Shomron

## Abstract

Anosmia is common in COVID-19 patients, lasting for weeks or months following recovery. The biological mechanism underlying olfactory deficiency in COVID-19 does not involve direct damage to nasal olfactory neurons, which do not express the proteins required for SARS-CoV-2 infection. A recent study suggested that anosmia results from downregulation of olfactory receptors. We hypothesized that anosmia in COVID-19 may also reflect SARS-CoV-2 infection-driven elevated expression of regulator of G protein signaling 2 (*RGS2*), a key regulator odorant receptor, thereby silencing their signaling. To test our hypothesis, we analyzed gene expression of nasopharyngeal swabs from SARS-CoV-2 positive patients and non-infected controls (two published RNA-sequencing datasets, 580 individuals). Our analysis found upregulated *RGS2* expression in SARS-CoV-2 positive patients (FC=14.5, Padj=1.69e-05 and FC=2.4; Padj=0.001, per dataset). Additionally, RGS2 expression was strongly correlated with PTGS2, IL1B, CXCL8, NAMPT and other inflammation markers with substantial upregulation in early infection. These observations suggest that upregulated expression of *RGS2* may underlie anosmia in COVID-19 patients. As a regulator of numerous G-protein coupled receptors, RGS2 may drive further neurological symptoms of COVID-19. Studies are required for clarifying the cellular mechanisms by which SARS-CoV-2 infection drives the upregulation of *RGS2* and other genes implicated in inflammation. Insights on these pathways may assist in understanding anosmia and additional neurological symptoms reported in COVID-19 patients.

## Introduction

Deficiency or complete loss of smell (anosmia) is common in COVID-19, affecting a substantial number of patients and sometimes lasting for weeks or months following recovery (1,2). Anosmia in COVID-19 patients was suggested as predictor for post-COVID-19 fatigue syndrome (“long COVID”; 3) and observed as a comorbidity with persistent post-COVID inflammation (4). The biological pathways underlying olfactory deficiency in COVID-19 remain unclear. Albeit SARS-CoV-2 may enter the brain (5,6), it does not seem to be capable of infecting olfactory neurons (7). It was recently shown that SARS-CoV-2 infection causes downregulation of olfactory receptors in olfactory neurons, which may explain COVID-19 related anosmia (8). We hypothesize that the olfactory dysfunction in COVID-19 may additionally reflect an infection-driven upregulation of *RGS2* (regulator of G protein signaling 2), a key regulator of nasal cavity G protein-coupled odorant receptors, whose signaling is diminished by the RGS2 protein (9). RGS2 is known to be co-expressed with odorant receptors, and injection of RGS2 antibody into olfactory neurons was shown to enhance calcium currents in olfactory neurons stimulated with odorants (10). Here, we present findings from analysis of RNA-sequencing data from nasopharyngeal samples, which show higher *RGS2* mRNA levels in COVID-19 patients. While the biological pathways upregulating *RGS2* mRNA in SARS-CoV-2 positive nasal tissues remain to be established, we present evidence that this is a consequence of the acute inflammation caused early in the span of infection. We present additional evidence for a correlation for higher RGS2 expression in populations with increased risk of COVID-19-induced anosmia.

## Methods

We analyzed RNA-sequencing data from two NCBI Gene Expression Omnibus (GEO) database: GSE163151 (11) and GSE152075 (12) to comparing the expression of *RGS2* in SARS-CoV-2 positive and negative nasopharyngeal epithelial cells. For this analysis we applied a generalized linear model under the standard DESeq2 method (13). Significant findings were reported for Padj<0.05. Additionally, Spearman correlation was calculated between genes for identifying genes with significant correlations with *RGS2* expression levels. Enrichment analysis was performed using Gene Ontology enRIchment anaLysis and visuaLizAtion tool (Gorilla) (14). Figures were plotted using GraphPad Prism version 9.3.1 for Windows, GraphPad Software, La Jolla, CA, USA.

## Results

We observed a 14.5-fold increase of *RGS2* mRNA expression in nasopharyngeal swabs of COVID-19 patients, compared with controls in both datasets (FDR=1.7e-05; **Figure 1A** and FC=2.4; Padj=0.001693; **Figure 1B**). The expression of RGS2 correlated with multiple, genes many of which were associated with the immune signaling and inflammation observed in COVID-19. *CXCL8* (C-X-C motif chemokine ligand 8), which codes for interleukin-8, had the strongest correlation with *RGS2* (R=0.91, P<3e-16 and R=0.83, P<3-16; **Figure 2**), followed by *PTGS2* (R=0.89, P<2.2e-16; R=0.8; P<2.2e-16; **Figure 3**), *NAMPT* (R=0.85, P<2.2e-16; R=0.79; P<2.2e-16; **Figure 4**) and *ILB1* (R=0.79, P<2.2e-16; R=0.71; P<2.2e-16; **Figure 5**)

**Figure 1.**
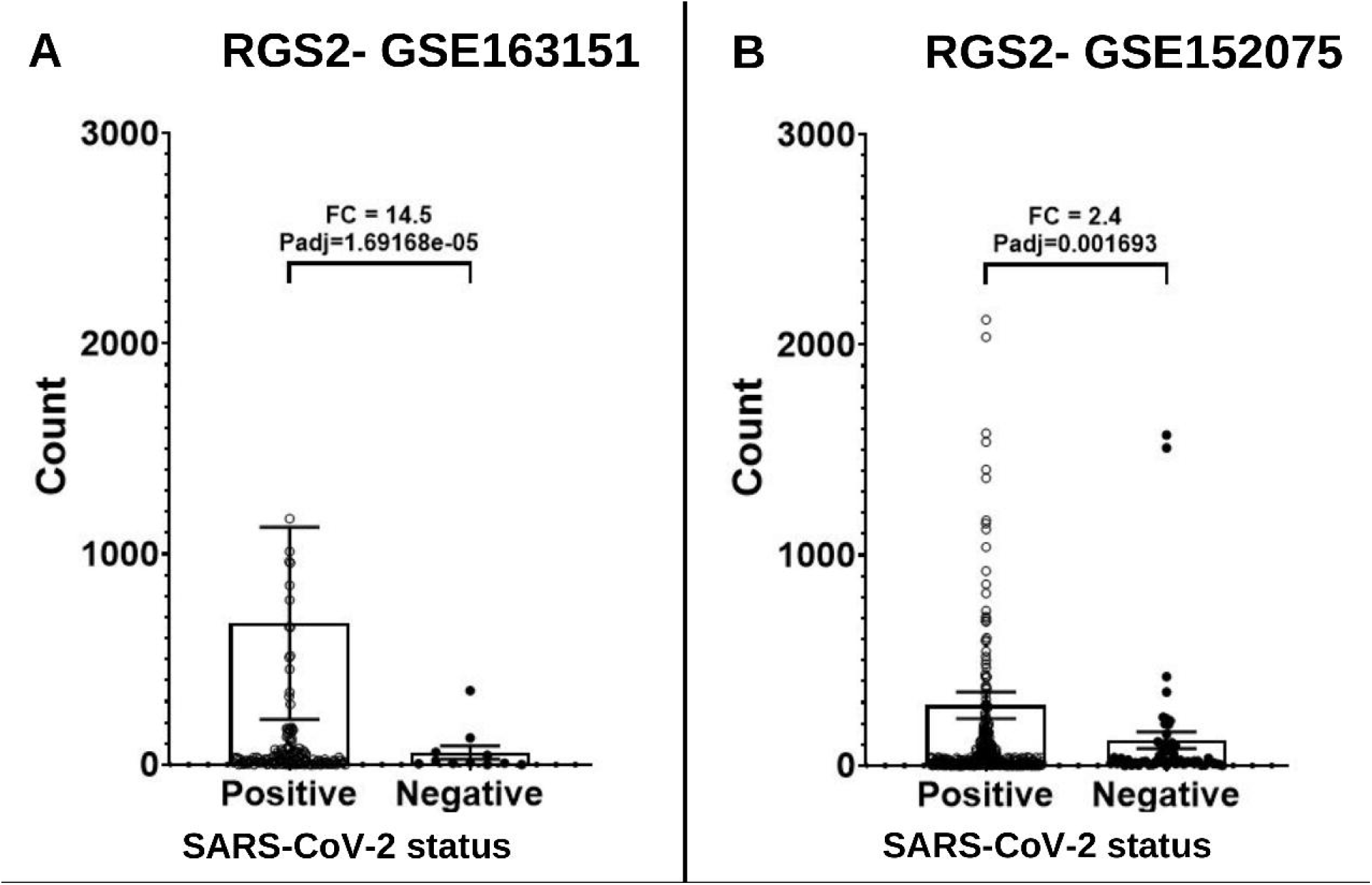
Expression levels of *RGS2* mRNA from RNA-sequencing of nasopharyngeal swabs of SARS-CoV-2 positive patients and negative controls. (A) Dataset GSE163151: higher expression of *RGS2* (FC= 14.5, padj=1.69e-5) in SARS-CoV-2 positive patients (138 samples) vs. negative controls (11 samples). (B) Dataset GSE152075: higher expression of *RGS2* (FC= 2.4, padj=0.0017) in SARS-CoV-2 positive patients (377 samples) vs. negative controls (54 samples). Samples with over 3000 *RGS2* counts (five and nine SARS-CoV-2 positive patients, respectively) are not shown due to scale limitations, but are included in the statistics.

**Figure 2.**
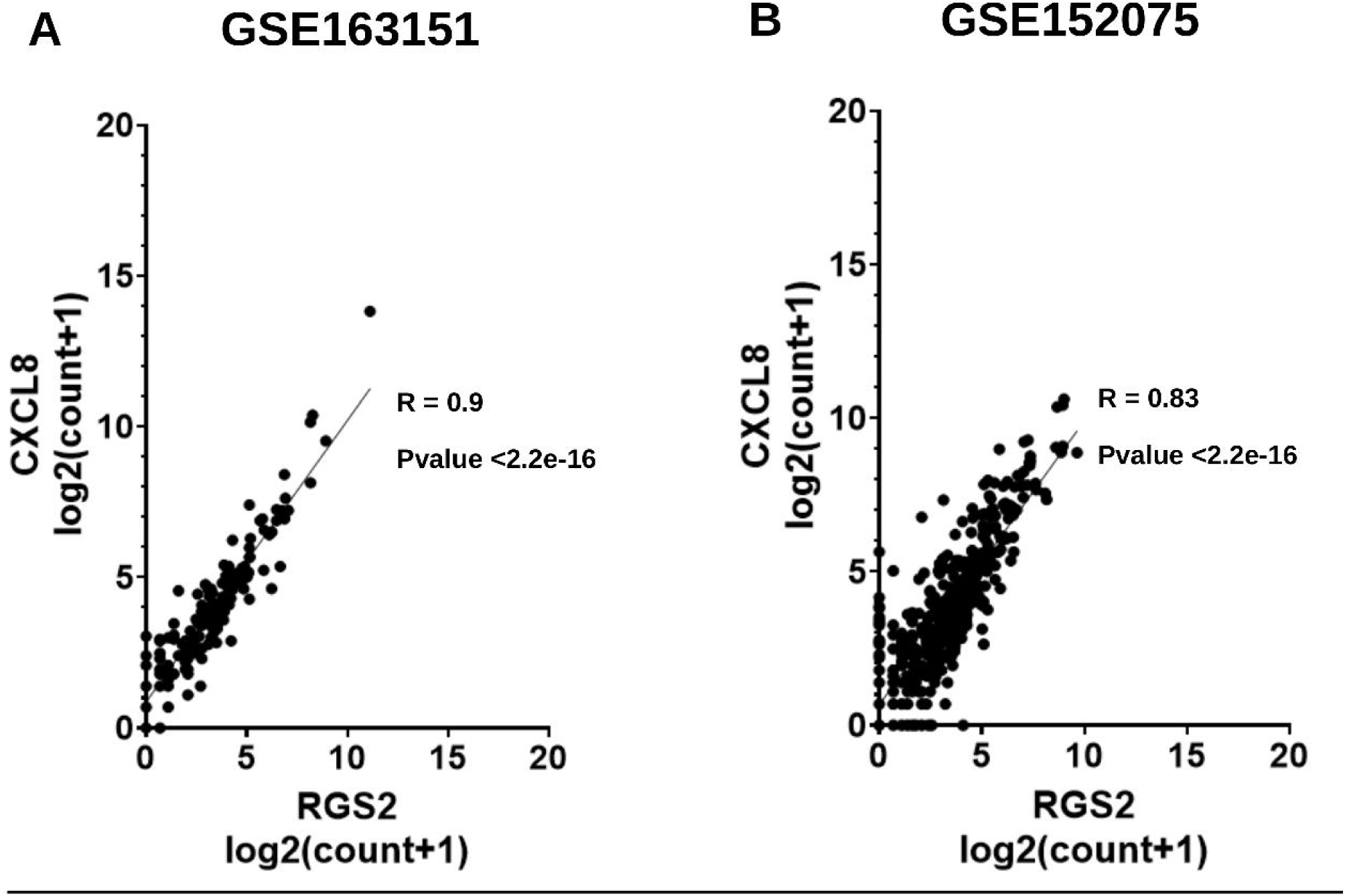
Correlation between expression levels of *RGS2* and *CXCL8* from RNA-sequencing of nasopharyngeal swabs. Spearman correlations are shown for (A) Dataset GSE163151; (B) Dataset GSE152075.

**Figure 3.**
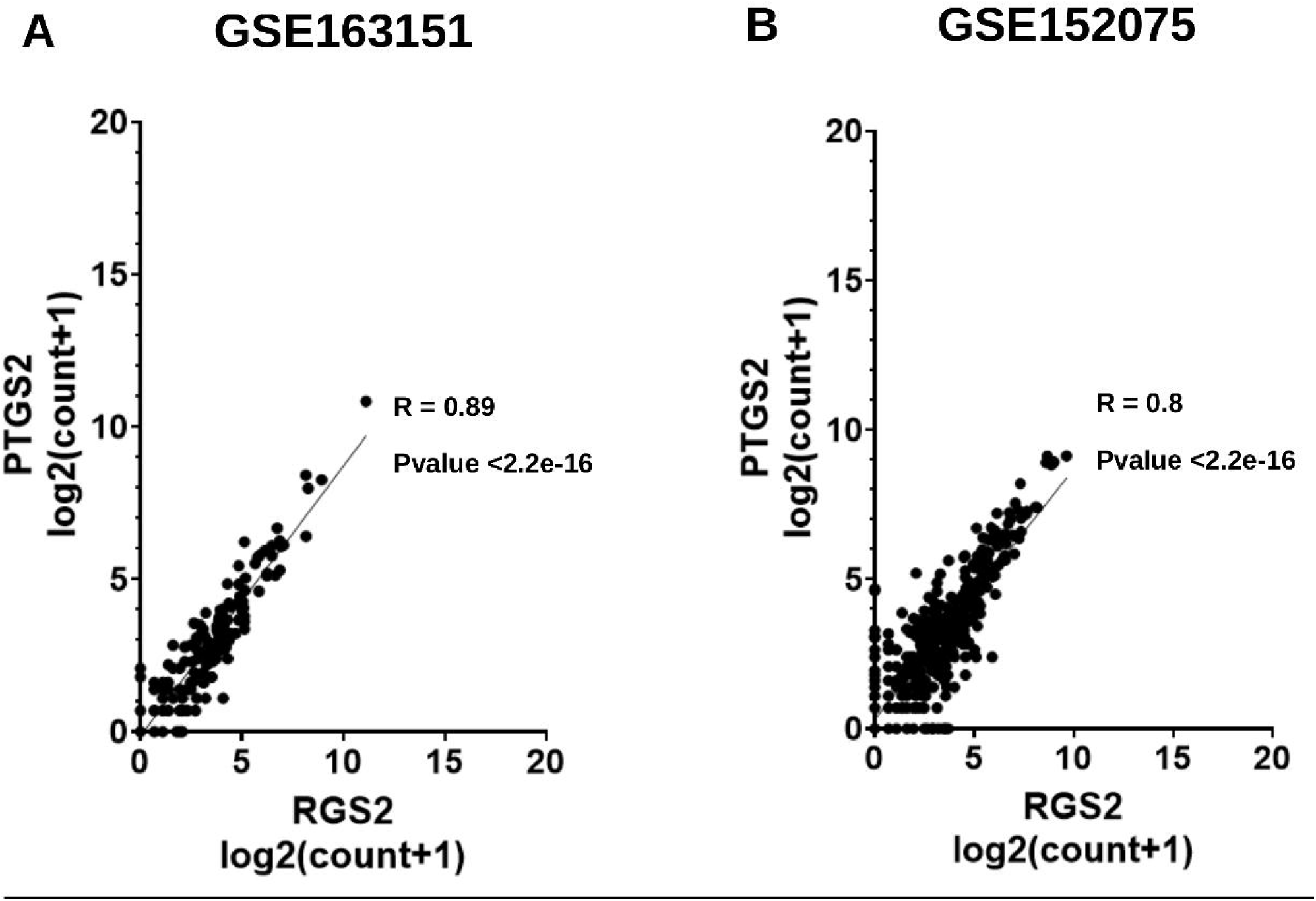
Correlation between expression levels of *RGS2* and *PTGS2* from RNA-sequencing of nasopharyngeal swabs. Spearman correlations are shown for (A) Dataset GSE163151; (B) Dataset GSE152075.

**Figure 4.**
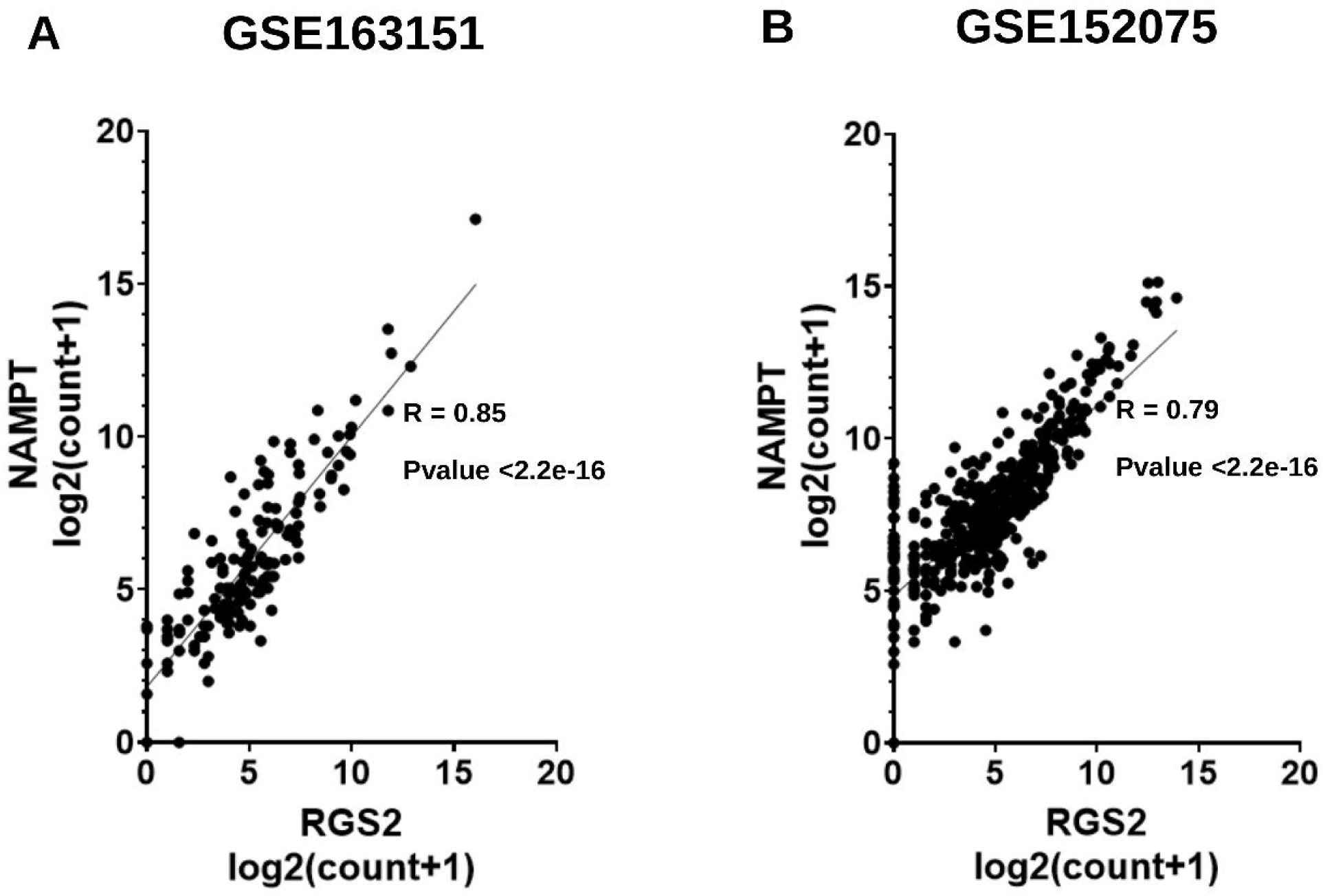
Correlation between expression levels of *RGS2* and *NAMPT* from RNA-sequencing of nasopharyngeal swabs. Spearman correlations are shown for (A) Dataset GSE163151; (B) Dataset GSE152075.

**Figure 5.**
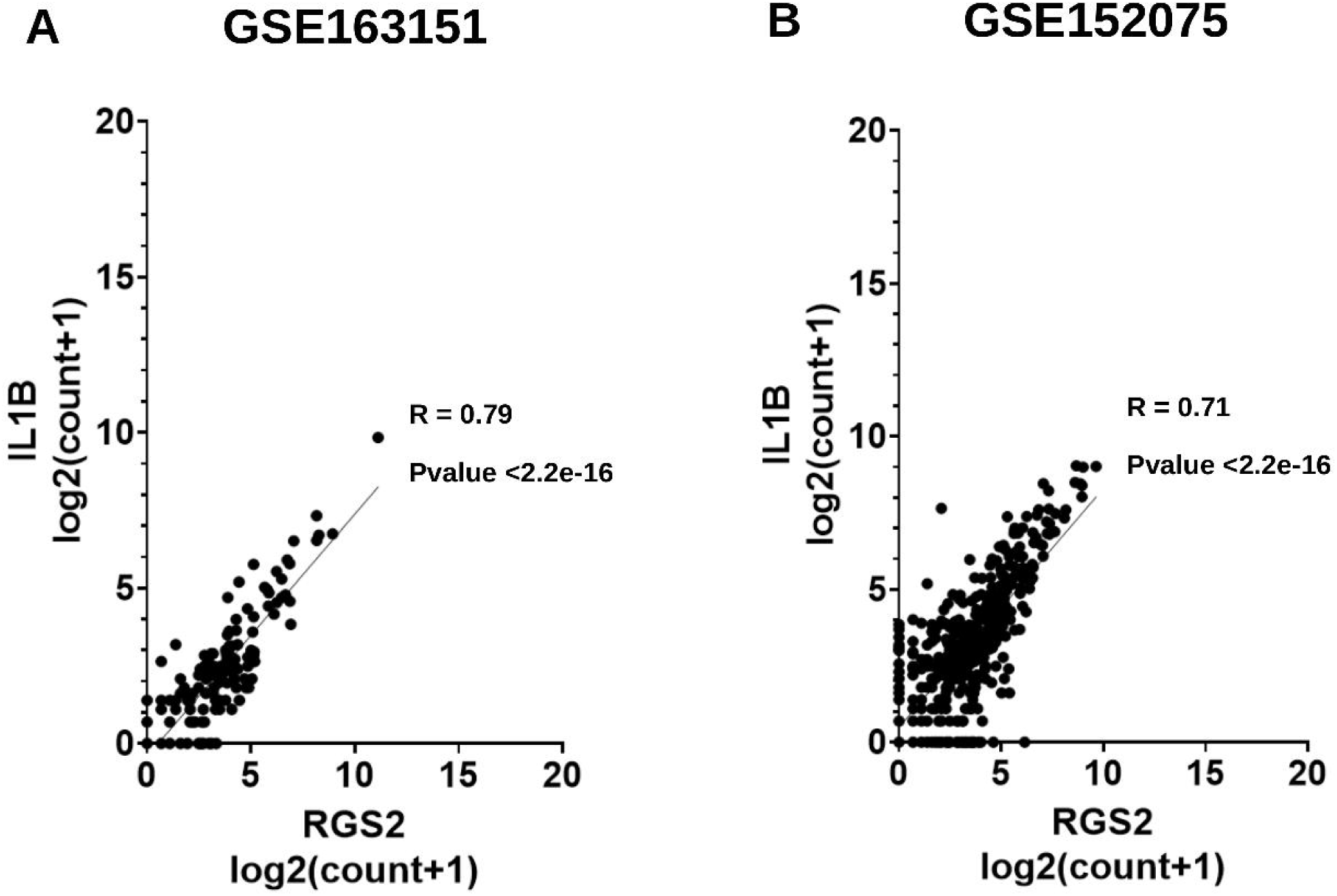
Correlation between expression levels of *RGS2* and *ILB1* from RNA-sequencing of nasopharyngeal swabs. Spearman correlations are shown for (A) Dataset GSE163151; (B) Dataset GSE152075.

The set of genes most correlated with *RGS2* expression (P<2e-16; **Table 1**) was significantly enriched for cellular response to interleukin-8, neutrophil aggregation and G protein-coupled receptor activity.

**Table 1.**
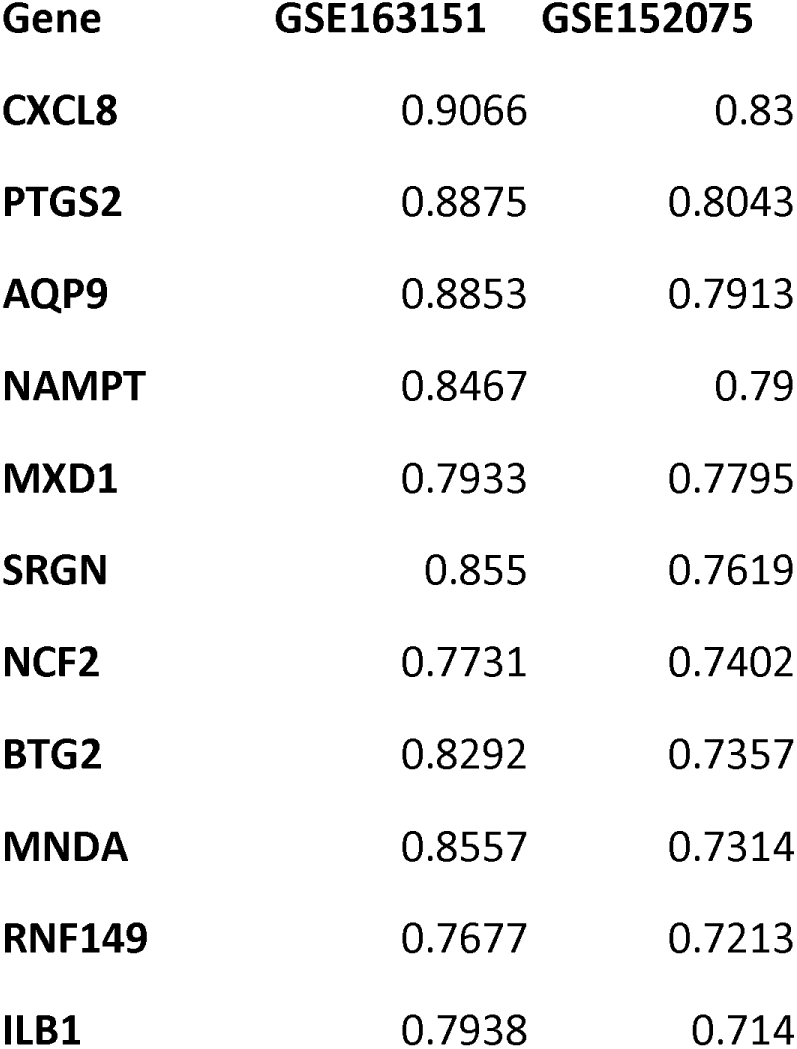
List of genes with expression levels correlations with *RGS2* in nasopharyngeal swabs from SARS-CoV-2 positive and negative individuals. Genes in GSE163151 and GSE152075 found with mRNA expression correlated with *RGS2* expression (with R>0.75 in at least of these GSE files) are listed by decreasing R values for GSE152075. All correlations had p<2.2e-16. See Methods for further information.

| **Gene** | **GSE163151** | **GSE152075** |
| --- | --- | --- |
| **CXCL8** | 0.9066 | 0.83 |
| **PTGS2** | 0.8875 | 0.8043 |
| **AQP9** | 0.8853 | 0.7913 |
| **NAMPT** | 0.8467 | 0.79 |
| **MXD1** | 0.7933 | 0.7795 |
| **SRGN** | 0.855 | 0.7619 |
| **NCF2** | 0.7731 | 0.7402 |
| **BTG2** | 0.8292 | 0.7357 |
| **MNDA** | 0.8557 | 0.7314 |
| **RNF149** | 0.7677 | 0.7213 |
| **ILB1** | 0.7938 | 0.714 |

## Discussion

We detected significant upregulation of *RGS2* mRNA expression in SARS-CoV-2 positive nasopharyngeal swabs, confirmed by two independent NCBI GEO datasets (**Figure 1**). RGS2 (regulator of G protein signaling 2) is recognized as the key regulator odorant receptor signaling (9). Moreover, RGS2 is co-expressed with nasal odorant receptors, and odorant receptor-mediated calcium currents were enhanced by injection of RGS2 antibody into olfactory neurons (10). Our findings therefore suggest that the elevated *RGS2* expression in SARS-CoV-2 positive nasopharyngeal cells is implicated in the common anosmia or reduced olfaction observed in many COVID-19 patients.

Anosmia is notable for being among the earliest symptoms to arise following SARS-CoV-2 infection (1-3, 15). *RGS2* expression is upregulated already during the first hours following SARS-CoV-2 infection, along with other early response genes, such as *PTGS2* (8,16).

COVID-19 associated anosmia is significantly more prevalent in young females (17), which are the demographic group with the highest *RGS2* blood expression (18). <u>Anosmia is most common among milder COVID-19 cases, which are also characterized as having higher levels of blood</u> *RGS2* expression compared to more severe patients (17,18).

We hypothesize that COVID-19 associated anosmia is caused by the strong, acute nasal inflammation elicited by interferon in mild COVID-19 cases (<u>19</u>). We suggest that *RGS2* upregulation is caused by inflammation of olfactory neurons, triggered via glial activation and IL-1-beta signaling, as previously observed in mouse models of intranasal lipopolysaccharide mediated inflammation (20).

A recent study of SARS-CoV-2 infection in model animals suggested that anosmia results from disruption of nuclear architecture (8). This study further corroborates the indirect inflammation hypothesis, detecting significant nasal IL-1-beta and RGS2 upregulation in the first hours of infection (8).

Another potential clue for the mechanism underlying COVID-19 associated anosmia could be derived from the recent emergence of the SARS-CoV-2 Omicron variant, which was accompanied by a drastic reduction of anosmia cases among British COVID-19 patients from 52.7% during the Delta wave to 16.7% during the Omicron wave (21). Consistent with our hypothesis, we postulate that the reduced immunogenicity of the Omicron variant leads to a significantly milder initial immune response (22) and in turn, milder induction of *RGS2* expression and thus reduced risk of anosmia. The relation between early viral load and anosmia was previously observed (23,24).

As a regulator of several G-protein coupled receptors, the elevated expression of RGS2 may drive further neurological symptoms observed in COVID-19 patients, sometimes lingering beyond negative SARS-CoV-2 PCR test findings (25). The highly correlated expression of *RGS2* with *CXCL8, PTGS2, NAMPT, ILB1* and further inflammatory genes in COVID-19 positive nasopharyngeal swabs (**Figures 1-5, Table 1**) suggests the involvement of a common regulator, which might explain the distinct classes of COVID-19 symptoms. Changes in the expression of circulating or exosomal microRNAs in COVID-19 is one factor that might explain at least some of the observed gene expression alterations (26, 27). Further studies of COVID-19 animal models are required for clarifying the cellular mechanisms by which SARS-CoV-2 infection drives this differential expression. Insights into the pathways driving anosmia in COVID-19 may improve our understanding of additional neurological indications reported in these patients (28).

## Data Availability

All data was obtained from the following publicly available Gene Expression Omnibus (GEO) datasets: GSE163151 and GSE152075.

https://www.ncbi.nlm.nih.gov/geo/query/acc.cgi?acc=GSE163151

https://www.ncbi.nlm.nih.gov/geo/query/acc.cgi?acc=GSE152075

## Abbreviations

ACE2: angiotensin-converting enzyme 2
COVID-19: coronavirus disease-2019
CXCL8: C-X-C motif chemokine ligand 8
GEO: gene expression omnibus
IL1B: interleukin 1 beta
NAD: nicotinamide adenine dinucleotide
NAMPT: nicotinamide phosphoribosyltransferase
NCBI: National Center for Biotechnology Information
PTGS2: prostaglandin-endoperoxide synthase 2
RGS2: regulator of G protein signaling 2
SARS-CoV-2: severe acute respiratory syndrome coronavirus 2
TMPRSS2: transmembrane serine protease 2

## Acknowledgements

The authors are grateful for Tal Zohar (Zotal Ltd., Israel) for his kind donation in support of this research project.

## Data Availability Statement

All data was obtained from the following publicly available Gene Expression Omnibus (GEO datasets: GSE163151 (https://www.ncbi.nlm.nih.gov/geo/query/acc.cgi?acc=GSE163151); and GSE152075 (https://www.ncbi.nlm.nih.gov/geo/query/acc.cgi?acc=GSE152075). See Methods for further information.

## Author Contributions

D. Gurwitz and N. Shomron conceived the study. E. Avnat and G. Shapira performed the datamining, analyzed the data and statistics and prepared the figures. All authors took part in writing the article.

